# Evaluation of the relationship between quantitative PCR results and cell culturing of SARS2-CoV with respect to symptoms onset and Viral load – a systematic review

**DOI:** 10.1101/2021.08.23.21262162

**Authors:** Rozenberg Gilad, Erster Oran, Ghersin Itai, Mandelboim Michal, Neuberger Ami, Schwartz Eli

## Abstract

**Background:** Viral culture is currently the most accurate method to demonstrate viability and infectivity of Severe acute respiratory syndrome Coronavirus (SARS-2 CoV). Routine clinical diagnosis, however, is mostly performed by PCR – based assays that do not discriminate between infectious and non-virus. Herein, we aimed to determine the correlation between positive viral cultures and either PCR positivity, the Cycle Threshold (Ct) or the number of viral copies.

**Methods:** A systematic electronic literature search was performed and studies that reported both viral SARS-CoV-2 culture and PCR–based assays were included. A separate search for samples from blood, urine, stool, breast milk and tears were performed. To convert Ct values reported in the reviewed studies were to viral genomic copies, calibration experiments with four different reaction performed, using quantified RNA molecules.

**Results:** A total 540 articles were reviewed, and 38 studies were included in this review. Out of 276 positive-culture of non-severe patients, 272 (98.55%) were negative ten days after symptoms onset, while PCR assays remained positive for up to 67 days. In severely ill or immunocompromised patients positive-culture was obtained up to 32 days and out of 168 cultures, 31 (18.45%) stayed positive after day 10. In non-severe patients, in Ct value greater than 30 only 10.8% were still culture-positive while in Ct >35 it was nearly universally negative. The minimal calculated number of viral genome copies in culture-positive sample was 2.5 × 10^3^ copies / mL. These findings were similar in immunocompromised patients. Recovering positive culture from non-respiratory samples was sporadically obtained in stool or urine samples. Conversion of Ct values to viral genome copies showed variability between different PCR assays and highlighted the need to standardize reports to correctly compare results obtained in different laboratories.

**Conclusion:** During the pandemic phase, non-severe COVID-19 patients who are recovering and are not immuno-suppressed, can be regarded as non-infectious, within 10 days from symptom onset, or with Ct value greater than 35 (or a calculated viral load lower than 1.2×10^3^ copies / mL). These findings have important implications for recovering patients and asymptomatic patients, with respect to isolation criteria. The conversion of Cq values to viral genome copies described herein may be useful in future work, enabling a more standardized comparison between results reported in different studies from different laboratories.

## Introduction

By the end of February 2021, coronavirus disease 2019 (COVID-19) has infected around 108 million people worldwide and caused more than 2.3 million deaths^1^.

In addition to its tragic medical implications, the COVID-19 outbreak also has considerable social and economic consequences^234^. The Organization for Economic Co-operation and Development (OECD) has forecast that the global economy will shrink by 6% in 2020^5^, the deepest recession since the Second World War^6^. Isolation and quarantine of infected people is one of the contributors to these consequences.

Viral culture is currently the most accurate method to demonstrate COVID-19 viability and infectivity^7^. However, it is not routinely performed due to its time-consuming, its labor-intensive nature, and the required safety measures for such viral pathogen propagation. Therefore, diagnosis of COVID-19 has largely relied on real-time reverse transcriptase polymerase chain reaction (rRT-PCR)^8^. This method is both rapid, highly sensitive and specific^7^. It can also provide quantitative data, under suitable conditions. However, RT-qPCR has a major limitation, as it might detect viral RNA of non-viable, and thus non-infective, viruses^9^. This phenomenon has been demonstrated in several viruses, such as Ebola virus^9^, respiratory syncytial virus^10^ and influenza^11^, as well as for SARS-CoV-2^12,13^. Recent studies using viral cultures deemed COVID-19 patients as non-infective after 5 to 7 days from onset of symptoms^12,14^. In contrast, the median duration of RNA shedding, as assessed by rRT-PCR, was 31 days in a cohort of Chinese patients^15^.

The RT-qPCR assay is based on a specific fluorescent signal that is generated when the target nucleic acid (viral RNA in the case of SARS-2) is present in the examined sample. The target sequence is amplified by PCR. The quantity of the target RNA is inverse correlated with the number of amplification cycles required to generate the positive signal. Thus, lower Ct levels represent a larger number of target nucleic acid copies in the examined sample^16^.

Despite the above-mentioned significant limitation, RT-qPCR results are usually reported in medical systems as a binary result (positive or negative). A positive result is frequently used to determine how long COVID19 patients need to remain isolated. Determining the isolation period of patients who have recovered from COVID-19 has immense medical, economical, psychological and social implications.

In this systematic review, we aimed to determine the correlation between COVID-19 infectivity, as expressed by positive viral cultures, in relation to the Ct thresholds and to number of viral copies which is a more accurate measurement, and in relation to time from symptoms onset.

## Methods

### Type of studies

We screened all clinical studies that included viral SARS-CoV-2 cultures taken from symptomatic and asymptomatic COVID19 patients of any age and described their positivity as a function of symptoms duration, cycle threshold (Ct) or number of viral copies determined by PCR. We have included randomized controlled trials (RCTs), non-randomized comparative cohorts and case series studies until August 15, 2020. We included viral cultures taken from the upper respiratory tract for the main analysis of outcomes, but also included studies that described cultures of materials from other sites (stool, urine, tears, breast milk) for further considerations. We also extracted the gene probes used in measuring the cycle threshold for *qPCR* positivity and methods of viral growth in culture.

#### Outcome measures

1. Duration of viral culture positivity in comparison to *qPCR* positivity as measured from the day of symptoms onset.
2. Duration of viral culture positivity in comparison to either Ct of a *qPCR* assay or to viral genome copies.
3. Ability to isolate SARS-CoV-2 from non-respiratory tract samples.

#### Search methods for identification of studies

We conducted a systematic electronic literature search with the PubMed and MedRxiv search engines. We used the following search terms: “COVID-19”\”coronavirus”\” SARS-CoV-2” combined with “culture”. We also used references of retrieved papers, including reviews or systematic reviews, to identify further studies. Two reviewers independently screened all studies published or appeared online as non-peer-reviewed pre-prints before August 1, 2020. We excluded all initially identified and retrieved articles that did not fulfill the inclusion criteria. In case of disagreement, a third reviewer acted as arbitrator.

##### Laboratory criteria for included studies

Nucleic acid testing: To be able to analyze the data presented in the included studies, they were selected so that either the PCR assays described by Corman et al^17^, or the SARS2 test developed by the US CDC^18^ were used. Additionally, a description of the sampling and RNA extraction procedures needed to be included, so that the conversion from Ct values to target copies could be performed.

##### Virus isolation

SARS-CoV-2 isolation was determined by the presence of a typical cytopathic effect and increased viral RNA in the supernatant.

## Conversion of Ct values to copy number and vice versa

Many of the studied reported Ct values without converting them to viral target copy number. In order to estimate the viral load in the samples described in these studies, we converted the Ct values reported, to viral genomic copies, based on the procedure performed in each study, as described herein.

RNA segments containing the amplified targets of E-Sarbeco, RdRp, N-Sarbeco and CDC N1 reactions were generated by *In vitro* transcription in the Israel Central Virology Laboratory (CVL). The RNA molecules were purified, quantified, and served as standards for the standard curve calibration. The number of target copies in 1 ng of purified RNA was calculated based on the amplified sequence length, thereby enabling the conversion from copy number to Ct values. Serial dilutions of each amplicon were examined by RT-qPCR to generate the standard curve for each reaction. The Ct values were then plotted against the calculated concentration using a semi-logarithmic scale graph, and the regression formula was deduced. The copy number per ml was then calculated for each report, based on its specific sampling and extraction method. Detailed description of the procedure is included in the Supplementary material of this review.

## Results

We reviewed 540 articles (figure 1). Finally, 14 studies were included in the comparisons between the duration of PCR positivity and viral culture positivity (Table 1A+1B). Twelve studies were included in the comparison between viral culture positivity and the cycle threshold and / or viral load of the molecular analyses (Table 2A+2B). Ten studies described viral cultures performed in specimens other than respiratory tract secretions (Table 3).

**Table 1.**
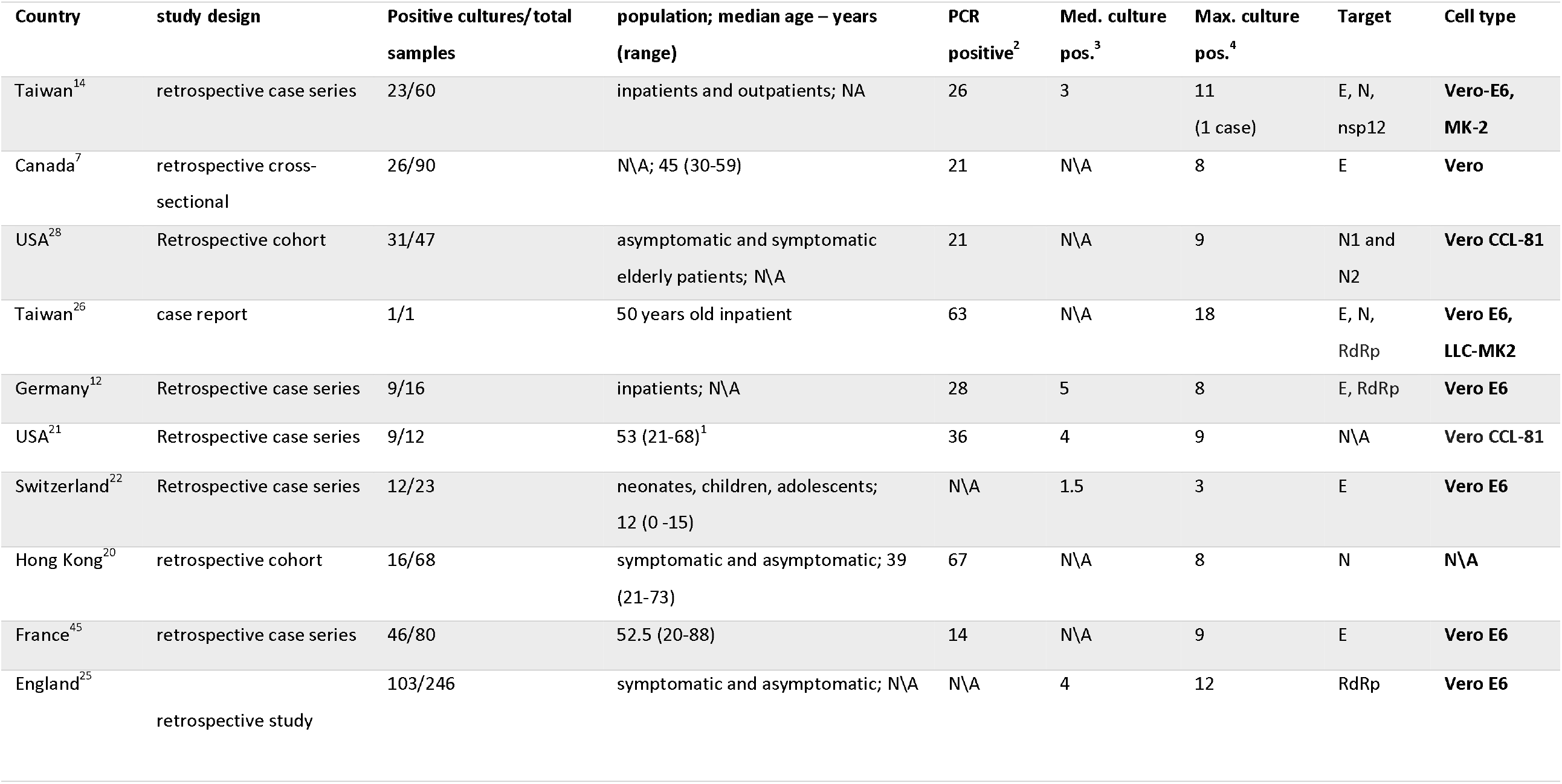
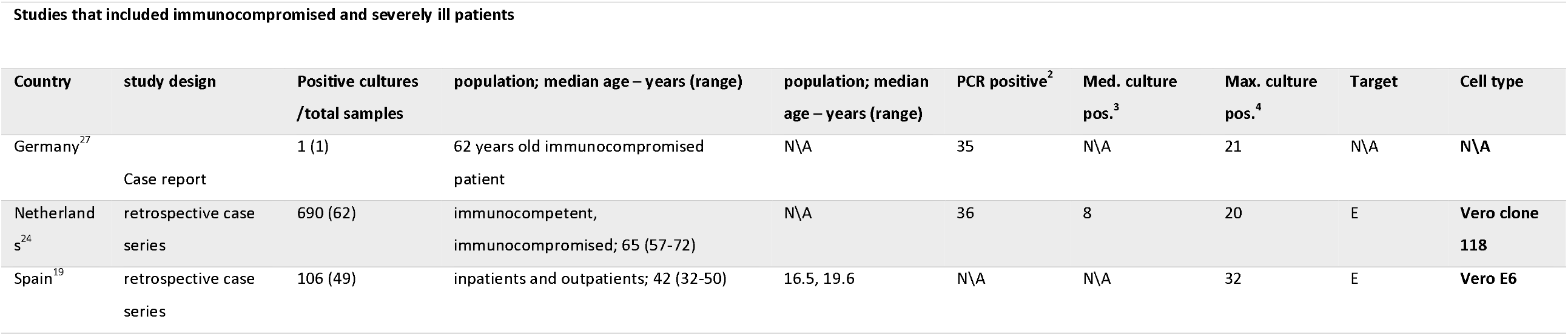
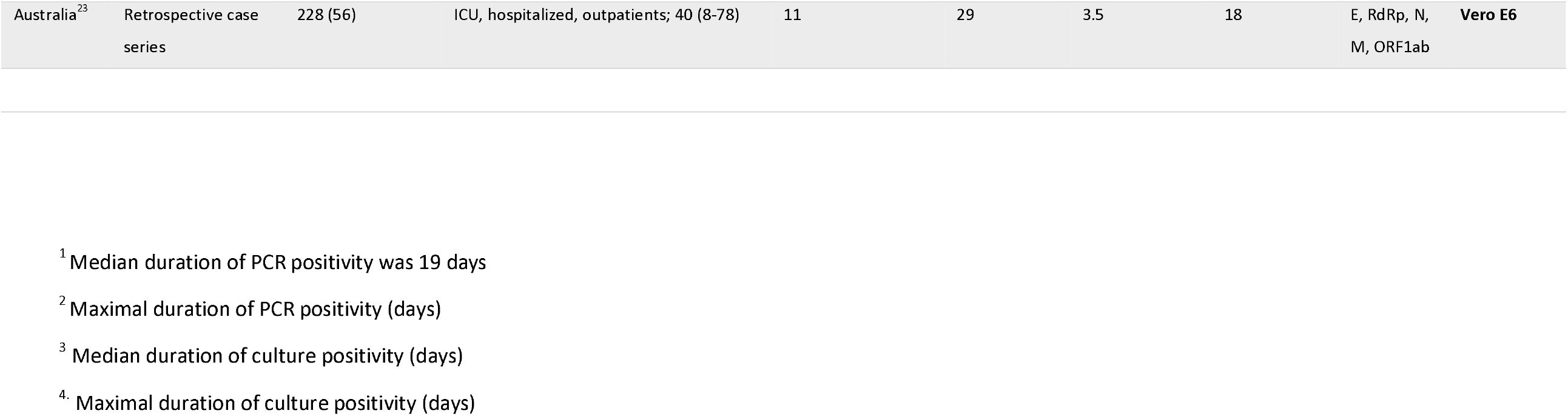
Comparison of the duration of SARS-CoV-2 PCR and culture positivity.

| Country | study design | Positive cultures/total samples | population; median age – years (range) | PCR positive<sup>2</sup> | Med. culture pos.<sup>3</sup> | Max. culture pos.<sup>4</sup> | Target | Cell type |
| --- | --- | --- | --- | --- | --- | --- | --- | --- |
| Taiwan<sup>14</sup> | retrospective case series | 23/60 | inpatients and outpatients; NA | 26 | 3 | 11<br>(1 case) | E, N,<br>nsp12 | Vero-E6,<br>MK-2 |
| Canada<sup>7</sup> | retrospective cross-sectional | 26/90 | N\A; 45 (30-59) | 21 | N\A | 8 | E | Vero |
| USA<sup>28</sup> | Retrospective cohort | 31/47 | asymptomatic and symptomatic elderly patients; N\A | 21 | N\A | 9 | N1 and<br>N2 | Vero CCL-81 |
| Taiwan<sup>26</sup> | case report | 1/1 | 50 years old inpatient | 63 | N\A | 18 | E, N,<br>RdRp | Vero E6,<br>LLC-MK2 |
| Germany<sup>12</sup> | Retrospective case series | 9/16 | inpatients; N\A | 28 | 5 | 8 | E, RdRp | Vero E6 |
| USA<sup>21</sup> | Retrospective case series | 9/12 | 53 (21-68)<sup>1</sup> | 36 | 4 | 9 | N\A | Vero CCL-81 |
| Switzerland<sup>22</sup> | Retrospective case series | 12/23 | neonates, children, adolescents; 12 (0-15) | N\A | 1.5 | 3 | E | Vero E6 |
| Hong Kong<sup>20</sup> | retrospective cohort | 16/68 | symptomatic and asymptomatic; 39 (21-73) | 67 | N\A | 8 | N | N\A |
| France<sup>45</sup> | retrospective case series | 46/80 | 52.5 (20-88) | 14 | N\A | 9 | E | Vero E6 |
| England<sup>25</sup> | retrospective study | 103/246 | symptomatic and asymptomatic; N\A | N\A | 4 | 12 | RdRp | Vero E6 |

**Studies that included immunocompromised and severely ill patients**
| Country | study design | Positive cultures<br>/total samples | population; median age – years (range) | population; median<br>age – years (range) | PCR positive<sup>2</sup> | Med. culture<br>pos.<sup>3</sup> | Max. culture<br>pos.<sup>4</sup> | Target | Cell type |
| --- | --- | --- | --- | --- | --- | --- | --- | --- | --- |
| Germany<sup>27</sup> | Case report | 1 (1) | 62 years old immunocompromised<br>patient | N\A | 35 | N\A | 21 | N\A | N\A |
| Netherlands<sup>24</sup> | retrospective case<br>series | 690 (62) | immunocompetent,<br>immunocompromised; 65 (57-72) | N\A | 36 | 8 | 20 | E | Vero clone<br>118 |
| Spain<sup>19</sup> | retrospective case<br>series | 106 (49) | inpatients and outpatients; 42 (32-50) | 16.5, 19.6 | N\A | N\A | 32 | E | Vero E6 |

|  |  |  |  |  |  |  |  |  |  |
| --- | --- | --- | --- | --- | --- | --- | --- | --- | --- |
| Australia<sup>23</sup> | Retrospective case series | 228 (56) | ICU, hospitalized, outpatients; 40 (8-78) | 11 | 29 | 3.5 | 18 | E, RdRp, N, M, ORF1ab | **Vero E6** |

<sup>1</sup> Median duration of PCR positivity was 19 days
<sup>2</sup> Maximal duration of PCR positivity (days)
<sup>3</sup> Median duration of culture positivity (days)
<sup>4</sup> Maximal duration of culture positivity (days)

**Table 2.**
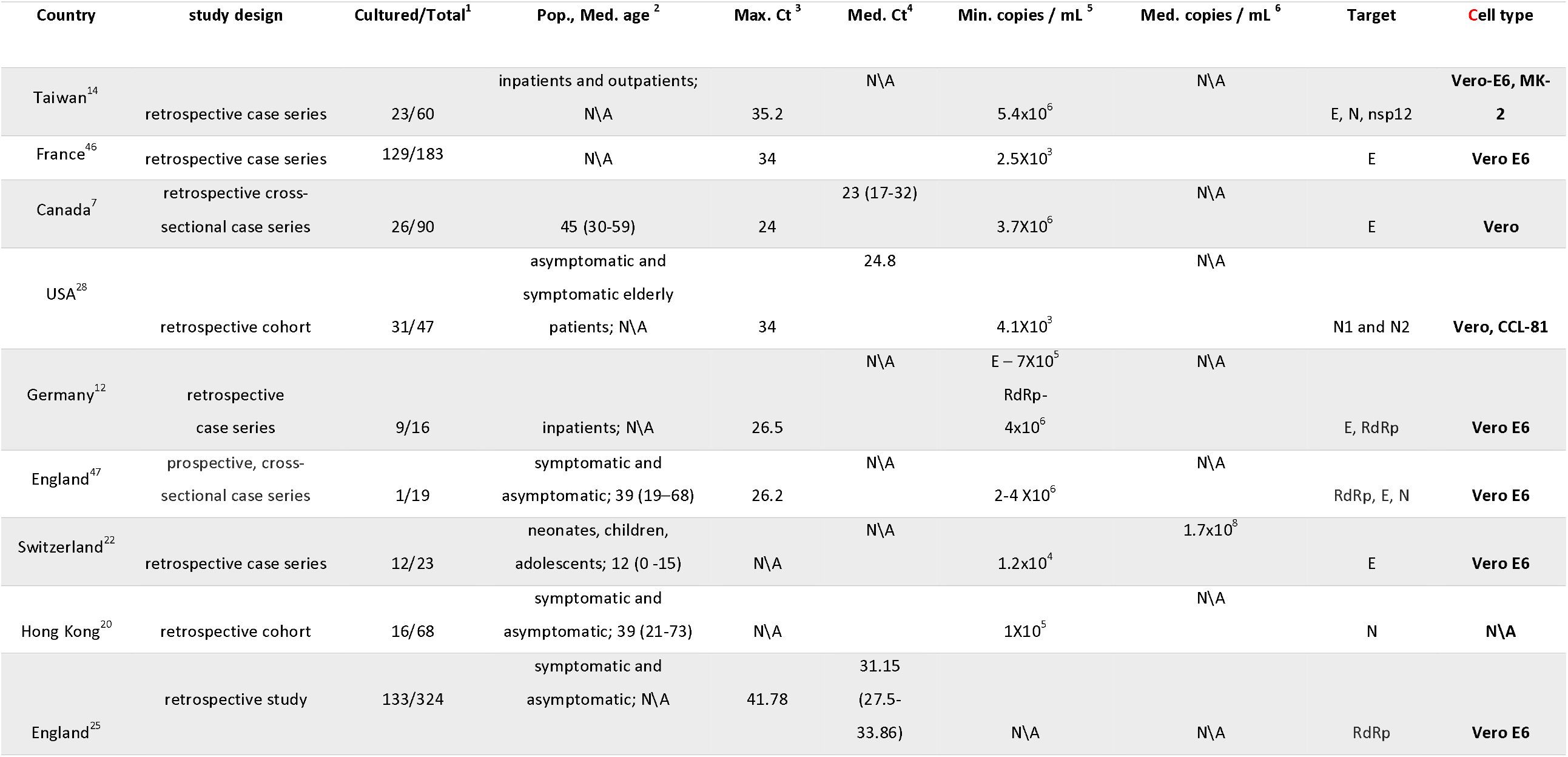
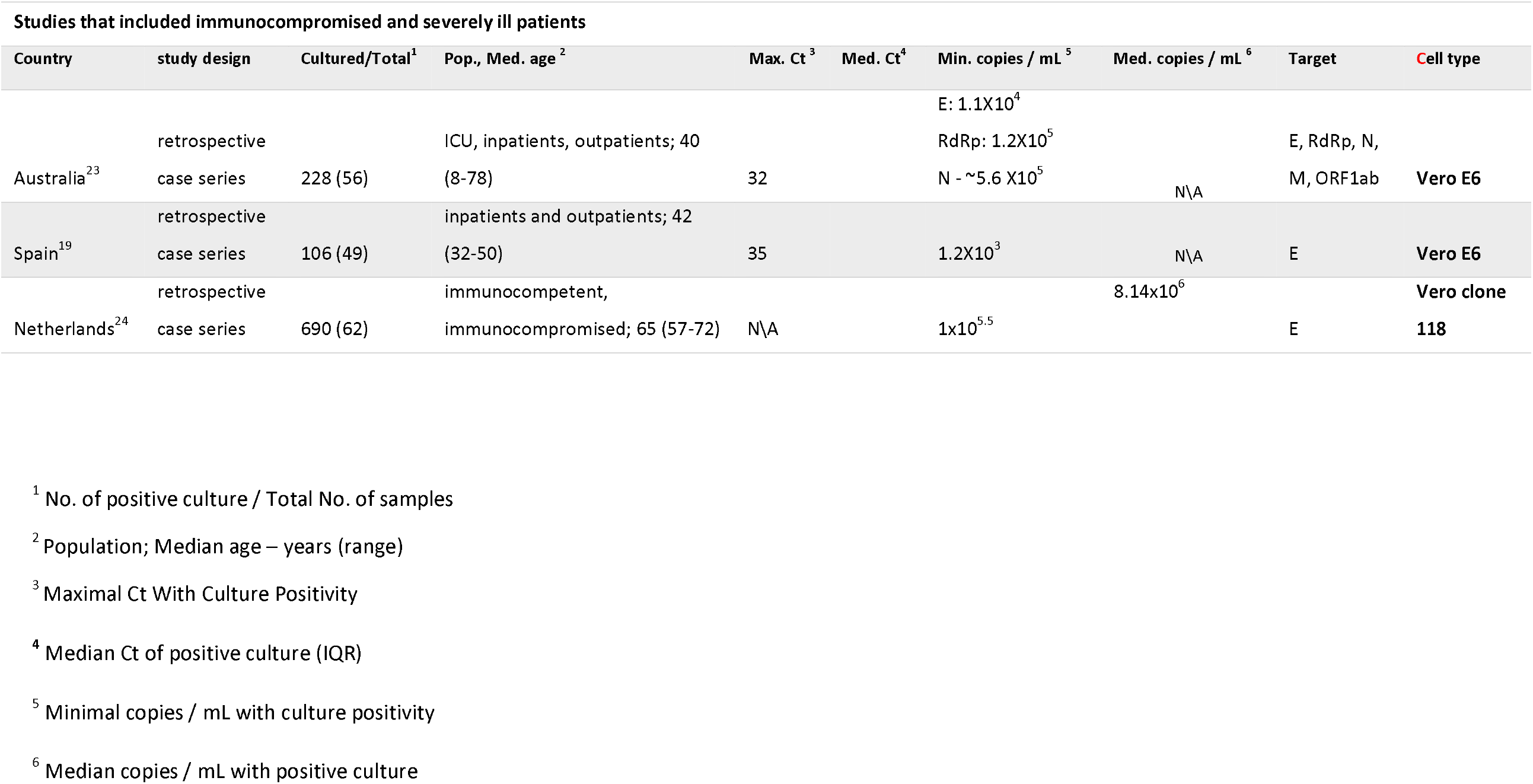
Minimal viral loads and maximal cycle threshold for positive SARS-CoV-2 cultures.

**Table 3.** Comparison of PCR positivity and culture positivity in non-respiratory body fluids.

| Body Fluid | Country | Study Design | Pos. culture/Pos. PCR<sup>1</sup> | Pop.; Med. age<sup>2</sup> | Target | Culture type |
| --- | --- | --- | --- | --- | --- | --- |
| **Blood** | United Kingdom<sup>29</sup> | Retrospective cohort | 0/16 | Acute and convalescent adult patients; 57 (46-76) | RdRp, ORF1ab | Vero E6 |
| **Blood** | USA<sup>30</sup> | Retrospective case series | 0/1 | 30-39 years old inpatient | N\A | Vero CCL-81 |
| **Stool** | USA<sup>21</sup> | Retrospective case series | 0/7 | 53 (21-68) | N\A | Vero CCL-81 |
| **Stool** | China<sup>30</sup> | Retrospective cohort | 2/4 | Inpatients; 44 (5-67) | ORF1ab | N\A |
| **Stool** | China<sup>31</sup> | Retrospective case series | 2/3 | Adult inpatients; N\A | ORF1ab | Vero E6 |
| **Stool** | Germany<sup>12</sup> | Retrospective case series | 0/4 | Adult inpatients; N\A | E, RdRp | Vero E6 |
| **Stool** | South Korea<sup>32</sup> | Retrospective case series | 0/3 | 63 (51-64); mild to critical disease | N\A | Vero |
| **Urine** | China<sup>33</sup> | Case report | 1/1 | 72 year old male inpatient with Severe COVID-19 | N\A | Vero E6 |
| **Urine** | South Korea<sup>32</sup> | Retrospective case series | 0/5 | 63 (51-64); mild to critical disease | N\A | Vero |
| **Breast Milk** | USA<sup>34</sup> | Prospective cohort | 0/1 | 25-29 years old woman | N\A | Vero E6 |
| **Tears** | Singapore<sup>35</sup> | Prospective study | 0/0 | Adult inpatients; N\A | N\A | Vero E6 |
| **Tears** | China<sup>36</sup> | Prospective case series | 0/2 | 54 (13-83) | N\A | **N\A** |

<sup>1</sup> No. of positive culture / Total No. of samples<sup>2</sup> Population; Median age – years (range)

**Figure 1:**
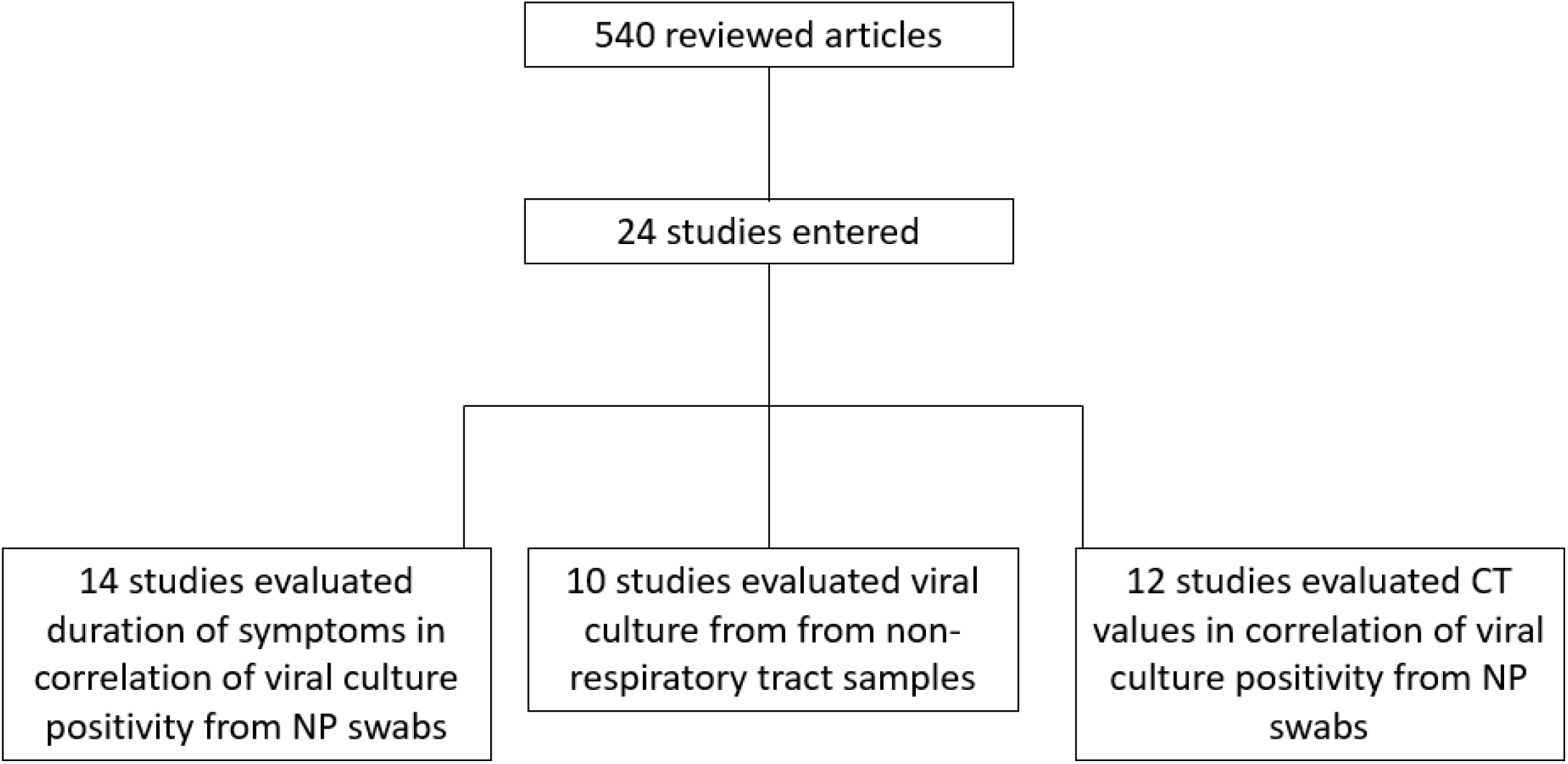

Examination of the duration of PCR and viral culture positivity from symptoms onset showed that out of 1668 positive PCR samples reported in the included studies, 444 (26.6%) cultures were positive, and 1224 (73.4%) were negative. Viral cultures were positive from between 6 days before the onset of symptoms, until 32 days after the onset of symptoms^19^. However, upon separating the immunocompetent-non-severe patients 272 out of 276 cultures (98.5%) were negative within 10 days of symptoms onset (Table 1A), while PCR assays remained positive for up to 67 days after the onset of symptoms^20^. In the seven studies that reported the duration of culture positivity in detail, the medians ranged between 1.5 and 8 days.^12,14,21,22,23,24,25^

From the group of studies that included immunocompetent patients with non-severe disease, cases positive after day 10 from symptoms onset included a case from Taiwan of a 50-year-old woman without any comorbidities and a clinically mild covid-19 had a positive viral culture by day 18^26^, another 2 samples from a series from England^25^, and one sample from second study from Taiwan^14^. Thus altogether, only 4 out of 276 (1.45%) samples stayed positive after day 10 of symptoms.

In case-series of severely ill-patients or immunocompromised patients, culture-positive samples were obtained up to 32 days of symptoms onset. For example, a recipient of a heart transplant, despite having mild disease, was reported to have positive viral cultures for 21 days^27^. In a work from the Netherlands that included immunocompromised patients, and more severely ill patients they had positive cultures up to 20 days from symptoms onset^24^. Finally, in a study from Spain, a severely ill patient had positive PCR assays for up to 32 days^19^. Altogether, for this group, 31 out of 168 positive samples (18.45%) stayed positive after day 10 of symptoms (Table 4). The summary of the 12 studies that examined the correlation between culture positivity and either cycle threshold (Ct) or viral load (VL) as expressed in genomic copies is presented in table 2.

**Table 4.**
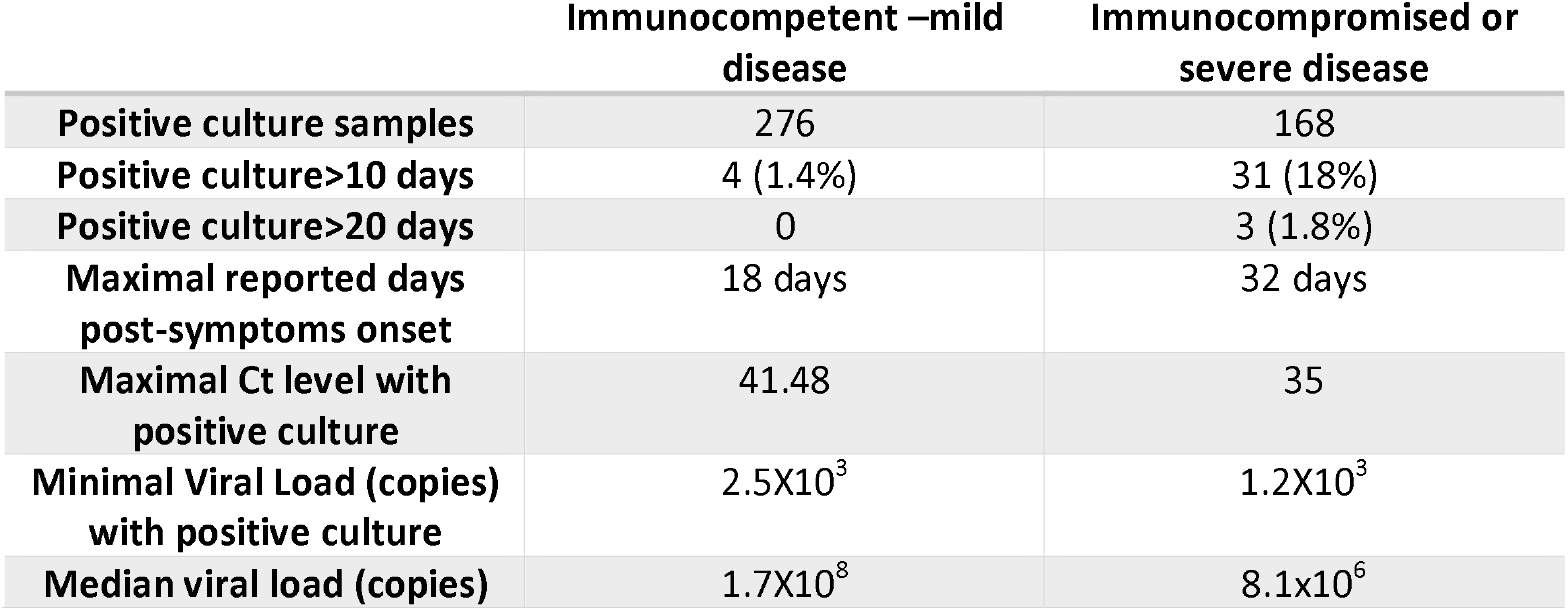
Positive culture Characteristics.

|  | **Immunocompetent –mild**<br>**disease** | **Immunocompromised or**<br>**severe disease** |
| --- | --- | --- |
| **Positive culture samples** | 276 | 168 |
| **Positive culture>10 days** | 4 (1.4%) | 31 (18%) |
| **Positive culture>20 days** | 0 | 3 (1.8%) |
| **Maximal reported days**<br>**post-symptoms onset** | 18 days | 32 days |
| **Maximal Ct level with**<br>**positive culture** | 41.48 | 35 |
| **Minimal Viral Load (copies)**<br>**with positive culture** | $2.5 \times 10^3$ | $1.2 \times 10^3$ |
| **Median viral load (copies)** | $1.7 \times 10^8$ | $8.1 \times 10^6$ |

The majority of samples had Ct level of <30, and only 59 samples out of 547 (10.8%) were culture-positive with Ct above 30. While positive samples with viable culture in Ct value above 35 were found only in 9 samples out of 547 (1.6%)^14,19,25^. In the five studies that reported the median Ct or VL in samples with positive cultures, Ct values range from 23, to the maximal median Ct was 31^25,7,28^. Median viral loads were 1.7×10^8^ and 8.14×10^6^. ^22,24^(Table 2)

In this regard, there were no major differences between immunocompetent and immunosuppressed or severely ill patients (Table 2 B). One exceptional case described a positive viral culture from a sample with a Ct of 41. No clinical data were provided for that specific patient regarding either immune status or illness severity.^25^

In addition to the information obtained from the literature, we added calculation of the viral genome copy number based on the Ct values. This was determined by generating standard curves for the E-sarbeco, N-sarbeco, RdRp and CDC-N1 reactions using diluted *In-vitro* transcribed RNA targets, as detailed in the Supplementary material file. The minimal calculated number of viral genome copies that were found in samples that were culture positive were 2.5 × 10^3^ and 1.2 × 10^3^ copies /mL in immunocompetent and immunosuppressed patients, respectively^19^. However, when studies where the median viral load is reported, there was a difference between the two population: 1.7×10^8^ copies /mL in immunocompetent patients and 8.14×10^6^ in immunosuppressed patients [Table 2]. A comparison of data among immunocompetent vs immunocompromised patients presented in Table 4.

Attempts to isolate the SARS-CoV-2 from non-respiratory tract samples are detailed in Table 3. No viral blood culture was positive in any patient with PCR-positive blood samples^21,29^. In studies of PCR-positive stool samples, stool viral cultures were positive in 4 out of 21 (19%) samples^12,21,30,31,32^. In one of the above-mentioned studies, no patient with culture –positive stools had diarrhea or any other GI symptoms^12^. A single case report describing a patient with PCR-positive and culture-positive urine was published^33^, in another study that tested urine specimens, none of the 5 samples were culture positive^32^. In a cohort of women with COVID-19, only one out of 64 samples of breast milk were PCR – positive, and none were culture positive^34^. Two studies tested tears in patients with or without conjunctivitis, in one study PCR and viral culture were negative for all samples^35^. In the second study two samples out of 60 yielded positive PCR results from a patient with conjunctivitis, non-yielded positive culture results^36^.

## Discussion

In this systematic review we analyzed data on COVID-19 viral culture positivity among patients with positive molecular tests for the virus. Among patients the were reported to be immunocompetent with mild clinical disease, nearly all patients (∼99%) are culture negative by day 10, even if PCR tests still were positive. In patients with severe COVID-19, or among immunocompromised hosts, viral cultures may remain positive for longer periods up to 32 days [table 1b].

In many viral infections including the coronaviruses (SARS-CoV, Middle East respiratory syndrome coronavirus) as well as in other RNA viruses such as influenza, Ebola and measles, it is well known that viral RNA can be detected long after infectious virus is cleared. This phenomenon is due to neutralization of the virus by the immune system. As viral RNA undergoes physical interaction with host cells and is protected by viral proteins, it can still be detected when the patient is no longer infectious. With measles virus for example, viral RNA can still be detected 6–8 weeks after the clearance of infectious virus^37,38,39^.

This *In-vitro* determination of the duration of infectivity correlates well with epidemiological studies. In a study from Taiwan, the authors describe a higher rate of infection in people exposed to index patients within 5 days of symptoms onset and lower rates of transmission after this time. In this study, the actual attack rate among 1818 patients whose exposure to index cases started within 5 days of symptom onset was much higher (1.0% [95% CI, 0.6%-1.6%]) when compared with those who were exposed later (0 cases from 852 contacts; 95% CI, 0%-0.4%)^40^. These epidemiological findings, supported by other similar observations, are consistent with our summary of *In-vitro* studies that showed nearly no viral infectivity beyond 10 days of symptoms onset^13^.

The other factor that may affect culture positivity is the viral load reflected either by a high number of viral genome copy number, or by a relatively low Ct count. In this study we found that practically all patients with a positive PCR test, but with Ct > 35 or with a calculated viral copies less than 1.2×10^3^ copies / mL, were culture negative. The reported result of a positive culture derived from a single clinical sample with a Ct value of 41^25^ is exceptional and warrants further investigation, in light of the accumulating data suggesting that samples with Ct values above 30-35 are non-infectious.

In this respect, it is important to consider the specific properties of the test performed. Different studies use different qPCR assays to evaluate the viral load in the human sample or in the cell culture. Although the vast majority of these tests have sufficient sensitivity to detect down to 50-100 copies in the reaction tube, they do vary, and the same sample can give a Ct result of 35 in one test, and Ct of 40 in another.

Furthermore, since the sensitivity of different PCR assays differs, and different laboratories use different extraction procedures (not including direct PCR tests, without RNA extraction), it is very challenging to accurately determine the actual viral load reported, based on Ct values. Ideally, results from quantitative PCR should be reported in viral RNA copies, rather than Ct values. However, accurate calculation of the viral load, as expressed by the number of target viral RNA copies detected in the sample, requires using calibrated standards. This is depended on availability of RNA standards in an established concentration and on performing a calibration test in parallel to the diagnostic test. This is not practical for many diagnostic laboratories, currently struggling with an overload of work. To accurately evaluate the viral load in the studies reviewed here, we undertook to experimentally determine the sensitivity of some of the qPCR assays described, as detailed in the Supplementary data. We show that while the E-Sarbeco and CDC N1 reactions have a similar sensitivity, the N-Sarbeco and RdRP reactions are approximately 5 to 10-fold less sensitive.

Adult patients with <u>severe</u> COVID-19 have prolonged viral shedding as reflected by a continuously positive PCR. A study from China showed that patients with severe disease had a median duration of viral shedding of 31.0 (IQR, 24.0–40.0) days from illness onset^15^. In a comparative observational study the median duration of viral shedding, reflected by positive PCR assays, was significantly longer among patients with severe disease (21 days, 14-30 days) than among patients with mild disease (14 days, 10-21 days; P=0.04)^41^. However, in none of these studies the presence of viable virus was studied. In this review, we present reports of samples from patients with either a severe disease or immunosuppression who were shedding infectious virus for longer than 10 days, up to 32 days after symptoms onset. Interestingly, the Ct levels and number of viral copies were similar in these cases to immunocompetent patients without severe disease. A possible explanation stems from the fact that the PCR test does not necessarily reflect infectivity; it provides information on the number of target copies present in the sample. Therefore, it is conceivable that in some immunocompromised patients, most or all of the detectable viral RNA is from infectious virus, while in other patients, most of the RNA is from partially degraded, non-infectious virions. In such cases, the PCR results are not consistent with actual infectivity and can be misleading. These assumptions are theoretical at this point and definitely warrant further investigation to allow comprehensive understanding of the infectivity state of such cases. Furthermore, persistent infection in immunocompromised patients can accelerate the development of new variants with increased virulence or infectivity, as suspected of the development of the recent British VOC^42^.

Interestingly in children, the rate of infectious virus isolation was largely comparable to that of adults, although two specimens yielded an isolate at a lower VL (1.2×10^4^ and 1.4×10^5^ copies/ml, respectively) than what was observed in adults^22^. Since SARS-CoV-2 shedding patterns in symptomatic children resemble those observed in adults, transmission of SARS-CoV-2 from children is plausible. The notion that small children are less infectious should therefore be attributed to other factors rather than the viral load per se^43^.

PCR-positive samples that were cultured successfully from non-respiratory tract such as: urine and stool, were reported in very few cases, thereby leading to the conclusion that infection via these secretion routes is probably negligible compared with respiratory route transmission.

No positive viral cultures of blood, tears and breast milk samples were reported thus far. Fecal-oral transmission, or transmission via contact with urine is therefore likely to be either non-existent or extremely rare. Similarly, as viable virus could not be detected in breast milk, breast feeding should be recommended for COVID-19 infected mothers who can take the necessary precautions^44^.

As the economic, social, and psychological toll of the COVID-19 pandemic becomes ever more evident, and second waves of infections inflicting many countries, it is vital to keep to a necessary minimum the number of days in which recovering COVID-19 patients remain isolated. From the data analyzed in this systematic review, it seems that for nearly all immuno-competent recovering patients with a non – severe infection, no more than ten days from the onset of symptoms would suffice. Such a recommendation would negate the need for additional PCR testing in such patients, as accepted now by several countries. The logistical, economic, social, and psychological benefits of such change are evident.

The maximal Ct values in a PCR – based assay that should be considered truly positive (rather than representing degrading genetic material) should be < 35 in nearly all cases. It is likely that for immunocompetent patients recovering clinically, a PCR test with Ct> 35 should be considered negative, leading to shortening of the recommended isolation periods. In the context of a local outbreak that strain laboratory resources and with limited isolation capabilities, a Ct value of 30 may also be considered, as only 10% of samples remained culture positive above this value.

A more standardized assessment of the infection state is calculating the viral genome copies, rather than relying on Ct values that are affected by the assay used. Reporting the test result using viral copy number, may minimize bias due to differences in assay performance, and allow comparison between results from different laboratories and different methods. This, however, requires the diagnostic laboratory to determine the reaction sensitivity and run routine calibration curves while performing clinical tests. In this study, we provide a schematic workflow for converting Cq values into target copies, allowing standardization of the method and comparison of results from different laboratories which use different qPCR tests.

The limitations of this review include lack of larger cohorts of COVID-19 with serial viral cultures, lack of standardization of laboratory techniques, and a reporting bias that probably leads to overestimation of cases with prolonged viral shedding. Another fundamental limitation is the need to draw practical conclusions from cell culture data. Due to obvious safety limitations, it is very complicated to perform SARS-CoV-2 infection experiments with animal model. Therefore, cell tissue culture (T/C) is the best practical alternative. However, inferring human viral infectivity based on T/C experiments needs to be done carefully, due to multiple stages, which the sample undergoes, before the test is performed. Sampling method, sampling media, transport timeline and storage conditions, may all affect the test results. Additionally, factors such as different susceptibility and presence/absence of effective immune response may also affect the quantity of infectious virus in the sample. A second limitation concerning T/C results is the different practices between different laboratories, different conditions of the cells used and different sample processing methods. Each of these factors separately can have a profound effect on the outcome. Combined, they render the comparison of different studies performed under different conditions very complicated. The reasons for unsuccessful culturing may stem from technical issues, and should therefore be interpreted carefully, when trying to correlate qPCR results with T/C infectivity. Nevertheless, as described above, the available epidemiological studies support the timeline delineated by culture positivity, suggesting that the infectious state is almost entirely during the early post-symptoms state.

In summary, the insights of this systematic review are important for policy makers worldwide. During the pandemic phase, symptomatic COVID-19 patients who are recovering from a mild infection and are not immuno-suppressed, can be regarded as non-infectious, within 10 days from symptom onset, with no additional testing needed. Based on the described findings, calculated viral load below 1.2×10^3^ copies per mL can be regarded as non-infectious. Recovering COVID-19 patients or asymptomatic patients whose tests yield Ct values greater than 35 may be regarded as non-infectious and may be freed from isolation. When the time of near elimination would (hopefully) come, more stringent criteria for defining non-infectiousness may be used. Urine, feces, blood, and breast milk may be regarded as largely non – infectious, based on the described results.

## Data Availability

All data is available in supplementary material

