## Supplemental material for "Evaluation of the relationship between quantitative PCR results and cell culturing of SARS2-CoV with respect to symptoms onset and Viral load – a systematic review"

Rozenberg Gilad, Erster Oran, Ghersin Itai, Mandelboim Michal, Neuberger Ami, Schwartz E.

**Supplementary material**

Although RT-qPCR is currently the gold standard for SARS2-CoV (and other pathogens) detection, most of the reports that describe diagnosis and study of the virus infection do not contain accurate quantification of the viral load, as can be inferred from the qPCR results. In order to analyze the studies described in this review, we generated calibration curves for the E, N and RdRp qPCR reactions that were described by Corman et al. (doi.org/10.2807/1560-7917.ES.2020.25.3.2000045) and are frequently used in many studies. We also performed similar analysis for the SARS-2-CoV N1 reaction that was developed and recommended by the US CDC (<https://www.cdc.gov/coronavirus/2019-ncov/lab/rt-pcr-panel-primer-probes.html> ), and is also used in some of the reports described herein. This allowed us to accurately convert the Cq values described in each such study, and compare the results of different analysis methods, which is very complicated to perform otherwise.

***Supplementary Materials and methods***

*Generation of RNA standards for the E, N and RdRp reactions*

DNA products corresponding to the target regions of the E, N and RdRp regions were amplified using primers that contained the minimal T7 promoter sequence in the 5’p end of the Fwd primer. This enabled *In vitro* transcription of the PCR prdoducts using the T7 Megascript kit (Fisher, ). The resulting RNA products were purified and their concentration was measured.

The RNA was aliquoted and stored at -80°C until further use.

*Generation of Standard calibration curves*

The RNA targets were diluted 10-fold and used in the corresponding RT-qPCR assays as described by Corman et al. (https://doi.org/10.2807/1560-7917.ES.2020.25.3.2000045) and by according to the CDC guidelins (<https://www.cdc.gov/coronavirus/2019-ncov/lab/virus-requests.html>).

**Results**

The Cq values obtained from the standard calibration RT-qPCR assays were plotted against the measured and calculated concentration of each RNA target. A regression formula was calculated from the semi-logarithmic correlation between the Cq values and the copy number for each target (Figure S1).

Based on the formula of each reaction, the conversion of Cq values to viral target copy number was calculated.

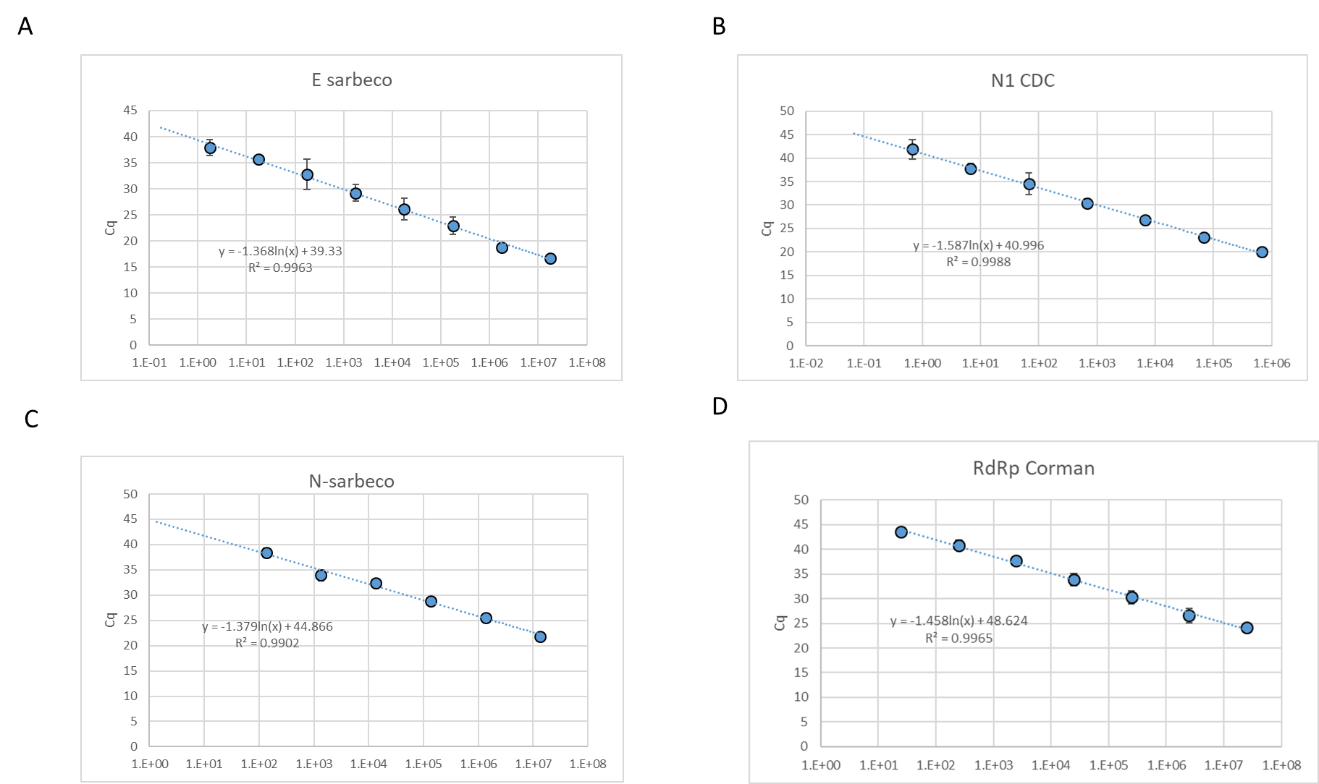
**Figure S1.** Calibration standards of the E-Sarbeco , RdRp, N-Sarbeco and CDC N1 reactions

The Cq values used to generate the calibration curves are detailed below in **Supplementary Tables S1 to S4**, for each target.

**Table S1.** Calibration assay for E-sarbeco reaction

| **E Sarbeco** | **Copies/RXN** | **average** | **stdev** |
| --- | --- | --- | --- |
| E(-3 | 1.76E+07 | 16.73 |  |
| E(-4 | 1.76E+06 | 18.83 | 0.39 |
| E(-5 | 1.76E+05 | 23.01 | 1.66 |
| E(-6 | 1.76E+04 | 26.13 | 2.11 |
| E(-8 | 1.76E+03 | 29.25 | 1.66 |
| E(-7 | 1.76E+02 | 32.76 | 2.89 |
| E(-9 | 1.76E+01 | 35.64 | 0.81 |
| E(-10 | 1.76E+00 | 37.90 | 1.49 |

**Table S2.** Calibration assay for CDC N1 reaction

| **N1 CDC** | **Copies/RXN** | **average** | **stdev** |
| --- | --- | --- | --- |
| nCOV-N1(-4) | 6.74E+05 | 20.10 | 0.007 |
| nCOV-N1(-5) | 6.74E+04 | 23.10 | 0.463 |
| nCOV-N1(-6) | 6.74E+03 | 26.90 | 0.548 |
| nCOV-N1(-7) | 6.74E+02 | 30.38 | 0.964 |
| nCOV-N1(-8) | 6.74E+01 | 34.52 | 2.253 |
| nCOV-N1(-9) | 6.74E+00 | 37.77 | 1.143 |
| nCOV-N1(-10) | 6.74E-01 | 41.87 | 2.126 |

**Table S3.** Calibration assay for N-sarbeco reaction

| **N-sarbeco** | **Copies/RXN** | **average** | **stdev** |
| --- | --- | --- | --- |
| N-sarbeco 10E-3 | 1.35E+07 | 21.87 | 0.55 |
| N-sarbeco 10E-4 | 1.35E+06 | 25.59 | 0.18 |
| N-sarbeco 10E-5 | 1.35E+05 | 28.84 | 0.25 |
| N-sarbeco 10E-6 | 1.35E+04 | 32.40 | 0.57 |
| N-sarbeco 10E-7 | 1.35E+03 | 33.92 | 1.07 |
| N-sarbeco 10E-8 | 1.35E+02 | 38.38 | 0.93 |

**Table S4.** Calibration assay for RdRP reaction

| **RdRp** | **Copies/RXN** | **average** | **stdev** |
| --- | --- | --- | --- |
| nCOV-RdRP(-3) | 2.48E+07 | 24.21439712 | 0.026529 |
| nCOV-RdRP(-4) | 2.48E+06 | 26.57239745 | 1.420449 |
| nCOV-RdRP(-5) | 2.48E+05 | 30.29038901 | 1.315342 |
| nCOV-RdRP(-6) | 2.48E+04 | 33.8583424 | 1.187385 |
| nCOV-RdRP(-7) | 2.48E+03 | 37.75137352 | 0.450568 |
| nCOV-RdRP(-8) | 2.48E+02 | 40.87861068 | 1.041671 |
| nCOV-RdRP(-9) | 2.48E+01 | 43.525 | 0.912168 |

Table S5 - Risk of bias and applicability judgments

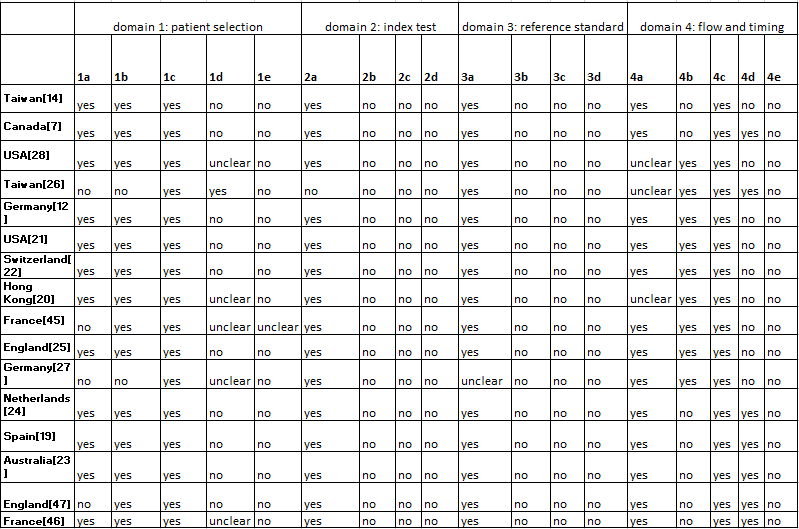

| **1a** | random sample of patients enrolled? |
| --- | --- |
| **1b** | a case-control design avoided? |
| **1c** | study avoid inappropriate exclusions? |
| **1d** | Could the selection of patients have introduced bias? |
| **1e** | Is there concern that the included patients do not match the review question? |
| **2a** | Were the index test results interpreted without knowledge of the results of the reference standard? |
| **2b** | If a threshold was used, was it pre-specified? |
| **2c** | Could the conduct or interpretation of the index test have introduced bias? |
| **2d** | Is there concern that the index test, its conduct, or interpretation differ from the review question? |
| **3a** | Is the reference standard likely to correctly classify the target condition? |
| **3b** | Were the reference standard results interpreted without knowledge of the results of the index test? |
| **3c** | Could the reference standard, its conduct, or its interpretation have introduced bias? |
| **3d** | Is there concern that the target condition as defined by the reference standard does not match the review question? |
| **4a** | Was there an appropriate interval between index test(s) and reference standard? |
| **4b** | Did all patients receive a reference standard? |
| **4c** | Did patients receive the same reference standard |
| **4d** | Were all patients included in the analysis? |
| **4e** | Could the patient flow have introduced bias? |
